# HIV–HPV Syndemic and Anal Precancerous Lesions Among MSM and Transgender Women in Pakistan: A Biological Continuum in High-Risk Sexual Networks

**DOI:** 10.64898/2026.05.28.26354356

**Authors:** Alyan Ahmed, Shaheer Hasan Rizvi, Ali Ahmed Rizvi, Fabeeha Ali, Aymen Haroon, Muslima Ejaz

## Abstract

**Background:** Sexual and gender minorities (SGM), including men who have sex with men (MSM) and transgender women, often face stigma, legal constraints, and limited access to sexual and reproductive health services. These conditions restrict prevention and care, increasing vulnerability to HIV and human papillomavirus (HPV) infections. While strong HIV–HPV interaction is documented in high-income settings, evidence from low- and middle-income countries remains limited. This study examines the burden, co-infection dynamics, and progression of HPV infection and anal dysplasia among MSM and transgender women in Pakistan.

**Methods:** A cross-sectional study was conducted between 15/09/2015 and 25/10/2016 among MSM and transgender women recruited from sexual health and antiretroviral therapy centers in Karachi. Eligible participants were aged ≥18 years and self-reported anal sex within the past 6 months (N=298). Two anal specimens were collected for HPV DNA detection and genotyping using PCR, and anal squamous intraepithelial lesions (ASIL) were assessed cytologically using the Bethesda classification. Associations were estimated using Cox proportional hazards regression algorithms to derive prevalence ratios (PRs).

**Results:** Among participants, 44% (n=133) were living with HIV. Overall HPV prevalence was 65.1%, rising to 87% among HIV-positive individuals compared to 48% among those without HIV (χ²p≤0.001). Likewise 28.9% of participants living with HIV were infected with two or more than two types of HPV as compared with 18.8% participants without HIV (χ²p≤0.001). HIV infection was strongly associated with HPV acquisition (adjusted PR 2.81, 95% CI 2.16–3.82). Among HPV-positive participants (n=194), 58.8% were co-infected with HIV. High-risk HPV was highly prevalent among those living with HIV (83.2% vs. 35.3% (χ²p≤0.001)), with HPV16 as the dominant oncogenic type. Multiple HPV infections were more common among HIV-positive individuals (χ²p≤0.001), and HIV seropositivity was 3.43 (95% CI: 2.55–3.51) times higher among those with high-risk HPV. Co-infected participants demonstrated prolonged smoking, longer duration of sex work, high-intensity sex work with limited condom negotiation, and higher prevalence of anal warts (all p<0.05). Anal dysplasia (ASIL) was present in 35% of participants and was higher among HIV-positive individuals (42.4% vs. 28.1%, p<0.001). HIV–HPV co-infection was independently associated with ASIL (adjusted PR 1.75, 95% CI 1.07–2.88), while high-risk HPV further amplified this risk (PR 3.04, 95% CI 1.75–5.26).

**Conclusion:** These findings demonstrate a biological continuum in HIV-positive MSM and transgender women, where HIV increases HPV acquisition, persistence, and multiplicity, accelerating progression to anal dysplasia. This reflects a syndemic shaped by biological interaction and structural vulnerability. Integrating HPV screening and vaccination within HIV services is essential to interrupt progression to cancer in this high-risk population.

**What this study adds:** This study provides context-specific evidence from Pakistan, a country experiencing a rapidly expanding HIV epidemic with low diagnosis and treatment coverage. It demonstrates a higher burden of high-risk and multiple HPV infections among individuals living with HIV and shows that HIV seropositivity was more than threefold higher among participants with high-risk HPV infection. These findings highlight a biological continuum linking HIV infection, HPV multiplicity, and anal dysplasia, and underscore the urgent need for integrated HIV–HPV prevention strategies in high-risk populations.

## INTRODUCTION

Human papillomavirus (HPV) is the most common sexually transmitted infection globally(1) and is typically acquired soon after sexual debut(2). While HPV infection is widespread, its burden is disproportionately higher among men who have sex with men (MSM) and transgender women, particularly in the presence of human immunodeficiency virus (HIV) infection(3, 4). Global estimates indicate that anal HPV prevalence approaches 90–93% among MSM living with HIV and remains substantially elevated even among HIV-negative MSM(5, 6). Persistent infection with high-risk HPV types, particularly HPV16 and HPV18, is the primary etiological factor for anal cancer and its precursor lesions, collectively termed anal squamous intraepithelial lesions (ASIL) (5, 7).

Although anal cancer is relatively rare in the general population, its incidence is markedly higher among MSM and is greatest among individuals living with HIV, with reported rates up to 70 times higher than those observed in the general population(5, 8, 9). This elevated risk is driven not only by behavioral exposures, such as receptive anal intercourse (10) and multiple sexual partnerships(11), but also by biological mechanisms linked to immunosuppression(12, 13). HIV infection impairs immune-mediated clearance of HPV, thereby facilitating viral persistence, multiplicity of infection, and progression to high-grade lesions and malignancy(14). Importantly, while antiretroviral therapy (ART) has substantially improved survival among people living with HIV, its impact on reducing HPV-associated dysplasia and anal cancer remains limited(15, 16).

These overlapping epidemics of HIV and HPV represent a classic syndemic, in which co-occurring infections interact synergistically within contexts of social and structural vulnerability (17, 18). This interaction is particularly pronounced in low- and middle-income countries (LMICs), where MSM and transgender women face intersecting barriers including stigma, discrimination, criminalization, and limited access to healthcare services, thereby restricting access to prevention, screening, and treatment (18, 19).

In Pakistan, the relevance of this syndemic is amplified by a rapidly evolving HIV epidemic. Recent estimates suggest that more than 200,000 individuals are living with HIV, with a substantial proportion remaining undiagnosed(20, 21). New infections have increased significantly over the past decade, with over 14,000 new cases reported in 2025 alone, reflecting ongoing transmission within key populations and urban sexual networks(20, 22). The epidemic remains concentrated among MSM, transgender women, and other marginalized groups, and is characterized by low testing coverage, delayed diagnosis, and suboptimal treatment uptake(22, 23). These dynamics create conditions for sustained co-infection with other sexually transmitted infections, including HPV, thereby increasing vulnerability to HPV-associated disease.(20, 24, 25).

MSM and transgender women in Pakistan represent socially marginalized populations with distinct identities, including male sex workers and hijras, individuals assigned male at birth who identify and live as women within a culturally specific gender system. Despite historical recognition, these groups experience significant social exclusion, economic vulnerability, and engagement in high-risk sexual networks, including commercial sex work. Limited condom negotiation power, barriers to healthcare access, and fear of stigma within healthcare settings further increase their vulnerability to sexually transmitted infections.

Anal squamous intraepithelial lesions (ASIL) are recognized precursor lesions for anal cancer, analogous to cervical intraepithelial neoplasia in cervical carcinogenesis. Studies from other settings have reported a high prevalence of abnormal anal cytology among MSM, particularly those living with HIV, highlighting the importance of early detection and intervention(26, 27).

In Pakistan, HPV vaccination has not yet been incorporated into routine immunization programs, and anal cancer screening services for high-risk populations remain largely unavailable. In the context of a growing HIV epidemic among MSM and transgender women, the absence of integrated HPV prevention strategies may further increase vulnerability to HPV-associated anal disease. Given the high burden of HIV–HPV co-infection and anal dysplasia reported globally among these populations, strengthening linkage between HIV care services and HPV preventive interventions, including screening and early detection of anal precancerous lesions, may represent an important public health opportunity in resource-constrained settings (28). Locally generated evidence is therefore essential to inform integrated prevention approaches tailored to key populations in Pakistan.

Against this backdrop, the present study adopts a syndemic framework to examine the interconnected burden of HIV infection, HPV infection, and anal dysplasia among MSM and transgender women in Pakistan.

The objectives of this study are to examine the relationship between HIV infection, HPV infection, and anal dysplasia among MSM and transgender women in Karachi, Pakistan, while characterizing HPV genotype distribution and the burden of anal precancerous lesions within this population. By generating locally grounded evidence, this study aims to inform integrated prevention strategies, including HPV vaccination, screening, and linkage with HIV care services, for this underserved and high-risk population.

## METHODS

### Study Design

This study was conducted as part of the quantitative phase of a multi-method research project and employed a cross-sectional design.

### Study Period

Data were collected between 15/09/2015 and 25/10/2016.

### Study Setting

The study was conducted in Karachi, Pakistan, the largest metropolitan city and capital of Sindh province. Karachi is a highly diverse urban setting with a population exceeding 14 million and represents a wide range of ethnic, socioeconomic, and cultural groups across Pakistan. Its demographic diversity makes it an appropriate setting for studying key populations at risk for sexually transmitted infections.

### Study Population and Recruitment

Participants included men who have sex with men (MSM) and transgender women aged 18 years or older who reported engaging in anal sex within the preceding six months.

Participants were recruited from two sites:

- **Setting I:** A public-sector antiretroviral therapy (ART) center at Civil Hospital Karachi, where transgender women living with HIV were approached during their scheduled ART visits.
- **Setting II:** A sexual health clinic operated by Perwaaz, a community-based organization providing services to sexual and gender minorities, including STI screening, HIV testing, and counseling.

Recruitment at the community-based site also utilized snowball sampling and peer referral strategies to reach hidden populations.

### Questionnaire Development

A structured, closed-ended questionnaire comprising 45 items was developed to capture information on sociodemographic characteristics, sexual behavior over the past six months, sexual history, knowledge related to sexually transmitted infections (STIs), and medical history relevant to HIV/AIDS and anal disease. The questionnaire was piloted among approximately 10% of the target population to ensure clarity, cultural appropriateness, and contextual relevance. Content validity was established through consultation with a panel of local experts. The average duration of each interview was approximately 35–40 minutes.

### Study Procedures and Data Collection

#### Questionnaire Administration

Participants were recruited on-site by a trained research team. After explanation of study objectives and procedures, written informed consent was obtained. For participants unable to read or write, a thumb impression was obtained. Face-to-face interviews were conducted in private settings, and participants were informed of their right to withdraw at any time without affecting their access to clinical services.

#### Blood Sample and Anal Swab Collection for HPV Testing and Cytology

Blood samples were collected from all participants for confirmation of HIV status, measurement of CD4+ T-cell counts, and assessment of viral load. Following collection, samples were transported to the laboratory and stored under controlled conditions until analysis.

Clinical examinations were conducted by trained physicians using an anal speculum to identify the presence of anal warts. Two anal swab specimens were collected from each participant: one for HPV DNA detection and the other for cytological assessment. The samples were placed in 3 mL of methanol-based fixative transport medium and stored at −70°C prior to laboratory processing(29). Cytological evaluation was performed by experienced cytopathologists.

### HPV DNA Extraction and Detection

#### DNA Extraction

DNA was extracted using the QIAamp DNA Mini Kit (Qiagen, Germany). Sample processing included centrifugation, resuspension in TE buffer, and enzymatic digestion using proteinase K. DNA was eluted and stored at −80°C until further analysis(29, 30).

#### HPV Detection and Genotyping

HPV DNA was detected using MY09/MY11 primers targeting the L1 region, resulting in amplification of a 450 base-pair fragment. A housekeeping gene (GAPDH) was used as a quality control measure, and samples negative for GAPDH were excluded. Nested multiplex PCR (NMPCR) was performed to identify HPV genotypes, including, low-risk types: HPV 6 and 11 and high-risk type; HPV 16, 18, 31, 33, 35, 52, 56, 58, and 59. Genotyping was performed using subtype-specific primers, and PCR products were analyzed through gel electrophoresis(30).

#### Cytological Analysis

Cytological analysis was performed using cytospin-prepared anal cytology slides stained with hematoxylin and eosin (H&E) and Papanicolaou stains. Cytological findings were classified according to the Bethesda system into; Atypical squamous cells of undetermined significance (ASCUS); low-grade squamous intraepithelial lesions (LSIL); atypical squamous cells - cannot exclude HSIL (ASC-H); high-grade squamous intraepithelial lesions (HSIL)(31)

### Study Variables

**Dependent variables** included the presence of any anal HPV infection, HPV16 infection, and abnormal anal cytology (ASIL), including ASCUS, ASC-H, LSIL, and HSIL.

**Independent Variables** were categorized into four domains: sociodemographic characteristics, sexual behavior, sexually transmitted infection (STI) knowledge, and clinical and biological factors. Key variables included HIV status, history of STIs, and the presence of anal symptoms. Behavioral measures comprised number of sexual partners, condom use, and sexual role, while lifestyle factors such as smoking and alcohol consumption were also assessed.

**Sample Size:** A minimum sample size of 267 participants was calculated for estimation of HPV prevalence, assuming an anticipated HPV prevalence of 50%, 6% precision, and a 5% level of significance. For the analysis of factors associated with HPV infection, a sample of at least 256 participants was estimated to provide 80% power to detect prevalence ratios (PRs) of 2.5 or greater across exposure frequencies ranging from 9% to 39%, at a 5% significance level. An additional 10% was added to account for inadequate biological samples, resulting in a target sample size of approximately 294 participants for HPV analyses.

For cytological assessment of anal squamous intraepithelial lesions (ASIL), a minimum sample size of 225 participants was required.

**Study Respondents:** A total of 320 participants were recruited. After excluding samples with inadequate biological material, 298 men who have sex with men (MSM) and transgender women were included in the HPV analysis. For cytological analyses, 27 samples (9.0%) were excluded due to insufficient biological material, resulting in a final analytical sample of 271 participants.

**Data Management and Statistical Analysis:** Data were entered, cleaned, and analyzed using the Statistical Package for Social Sciences (SPSS) version 24.0 (IBM Corp., Armonk, NY, USA).

Descriptive statistics were used to summarize participant characteristics. Continuous variables were presented as means with standard deviations (SD) for normally distributed data and medians with interquartile ranges (IQR) for non-normally distributed data. Independent sample t-tests were used to compare means between groups. Categorical variables were summarized using frequencies and proportions and compared using Pearson’s χ² test or Fisher’s exact test, as appropriate.

**HPV Prevalence and Genotype Analysis:** The overall prevalence of HPV infection, as determined by PCR, was calculated as the proportion of HPV-positive participants among all included individuals. Type-specific HPV prevalence was calculated as the proportion of participants positive for each HPV genotype among those who tested positive for HPV DNA.

Confidence intervals (95% CI) for prevalence estimates were calculated using the binomial distribution. Prevalence of “any anal HPV infection” and “any high-risk (HR) HPV infection” was further stratified by HIV status and sexual orientation, and comparisons between groups were conducted using Pearson’s χ² test.

To account for multiple comparisons across sexual orientation groups stratified by HIV status, Bonferroni correction was applied, with an adjusted significance threshold of p = 0.0083 (0.05/6).

**Cytological Outcome Analysis (ASIL):** The prevalence of anal squamous intraepithelial lesions (ASIL) was calculated as the proportion of participants with any abnormal cytology among all individuals included in cytological analyses. Corresponding 95% confidence intervals were estimated using the exact binomial method.

**Risk Factor Analyses:** Given the cross-sectional study design and relatively high prevalence of outcomes, Cox proportional hazards regression models with robust standard errors were used to estimate crude and adjusted prevalence ratios (PRs) and their 95% confidence intervals. For HPV-related analyses, the primary outcomes were; 1) Any anal HPV infection and 2) HPV16 infection. For cytological analyses, the primary outcome was; Presence of any ASIL (LSIL, ASCUS, ASC-H, HSIL)

Independent variables included biological and behavioral factors. Biological variables comprised HIV status, presence of anal sexually transmitted infections (STIs) including gonorrhoea (*Neisseria gonorrhoeae*), trichomonas infection, anal warts, and HPV infection status. Anal symptoms such as bleeding, itching, and discharge were also included.

Behavioral variables included number of sexual partners, condom use (consistent vs inconsistent), smoking status (including intensity and duration), and alcohol consumption (frequency categories).

Variables with p<0.20 in univariate analysis and those of biological or clinical relevance were included in multivariable models. A backward elimination approach was used to derive the final model. Robust standard errors were applied to obtain accurate confidence intervals.

Additionally, an independent samples t-test was conducted to compare the mean number of HPV genotypes between participants with and without ASIL.

All statistical tests were two-sided, and a p-value <0.05 was considered statistically significant, except where Bonferroni correction was applied.

### Ethical Approval

The study was approved by the Human Research Ethics Committee of The Aga Khan University (3612-CHS-ERC-15) and Dow University of Health Sciences Karachi Pakistan (IRB-557/DUHS/APPROVAL/2015/84). Written informed consent was obtained from all participants prior to enrollment.

## RESULTS

### Participant Characteristics

A total of 298 men who have sex with men (MSM) and transgender women were included in the HPV analysis. For cytological analyses, 271 participants were included after excluding 27 individuals (9.0%) due to insufficient biological material.

The mean age of participants was 28.8 years (±8.1), indicating a relatively young study population. Overall, 33.2% of participants were aged <25 years, 31.0% were aged 25–29 years, and 22.5% were aged ≥35 years. The majority were unmarried (77.9%), while 19.6% were married and 2.5% were separated or divorced. In terms of sexual identity, 57.6% identified as homosexual, 21.4% as bisexual, and 21.0% as transgender women. (Table 1)

**Table 1:** Sociodemographic, Behavioral and Clinical Characteristics Relevant to HPV Risk among MSM and Transgender Women (N = 298)

| Domain | Variable | n (%) / Mean ± SD |
| --- | --- | --- |
| Demographics | Age (mean ± SD) | 28.8 ± 8.1 |
|  | Age <30 years | 192 (64.4%) |
| | Education (≤ primary) | 119 (40.0%) |
|  | Unmarried | 234 (78.5%) |
| Sexual Identity | MSM | 167 (56.0%) |
|  | Transgender women | 63 (21.2%) |
|  | Bisexual | 68 (22.8%) |
| Sex Work & Exposure | Male sex workers | 237 (79.5%) |
| | Duration of sex work (years) | 12.4 ± 8.2 |
| | ≥50 clients (last 6 months) | 210 (70.5%) |
| Sexual Practices | Receptive anal sex | 264 (88.6%) |
|  | Inconsistent/No condom use | 226 (75.8%) |
| | Age at sexual debut (years) | 16.4 ± 4.7 |
| Substance Use | Smoking | 232 (77.9%) |
|  | Cigarettes <10/day | 128 (55.2%) |
|  | Alcohol use | 71 (23.8%) |
|  | Other substance use (any) | 154 (51.7%) |
|  | Hashish/Chars/Naswar | 101 (65.5%) |
| STI Burden | Other STIs | 167 (56.0%) |
|  | Genital warts (past 6 months) | 205 (68.8%) |
| HIV (Key Subgroup) | HIV positive | 131 (44.0%) |
| | Years since diagnosis | 3 ± 2 |
| | CD4 >350 cells/μl | 98 (74.8%) |
|  | Viral suppression | 100 (76.3%) |
|  | On cART | 117 (89.3%) |

Values are presented as mean ( ± SD) or percentages as appropriate

### HIV Status and Clinical Profile

Among all participants, 44% (n=131) were living with HIV. Among those living with HIV, 89.3% were receiving antiretroviral therapy (ART), 74.8% had CD4 counts greater than 350 cells/μL, and 76.3% had an undetectable viral load. Participants living with HIV were significantly older than HIV-negative participants (30.9 vs. 27.0 years, p<0.001). (Table 1)

### Sexual Behavior and Network Characteristics

A substantial proportion of participants were engaged in high-risk sexual networks, with 79.3% reporting involvement in sex work. The mean age at sexual debut was 16.4 years (±4.7), and the mean duration of sex work was 12.4 years (±8.2). High levels of partner turnover were observed, with 40.6% reporting more than 100 anal receptive partners in the past six months.

Sexual practices were predominantly high-risk, with 89.7% reporting primarily receptive anal intercourse. Condom use was suboptimal, with 60.1% reporting inconsistent use and 17.7% reporting never using condoms. Participants living with HIV reported a higher number of sexual partners, were more likely to engage in receptive anal intercourse, and had significantly lower condom use compared to HIV-negative participants. (Table 1)

### Substance Use and STI Profile

A high proportion of participants (77.5%) reported current smoking. The mean age of smoking initiation was 16.5 years, and the mean duration of smoking was 12.4 years. Alcohol use was less common, with approximately 20–28% of participants reporting alcohol consumption.

A substantial burden of sexually transmitted infections was observed, with 59.0% reporting a history of STIs and 68.8% reporting anal warts in the past six months, indicating a high prevalence of concurrent infections within the study population. (Table 1)

### Prevalence of HPV DNA, High-Risk HPV, and Genotype Distribution

The overall prevalence of anal HPV infection was 65.1% (194/298). HPV prevalence was significantly higher among participants living with HIV compared to those without HIV (87% vs. 48%, p≤0.001). Among HPV-positive individuals, 58.8% were co-infected with HIV, indicating a substantial overlap between the two infections.

High-risk HPV infection was markedly more prevalent among individuals living with HIV compared to HIV-negative participants (83.2% vs. 35.3%). Multiple HPV infections were also common among HIV-positive participants, affecting 68.0% of individuals. HPV16 was the most frequently detected oncogenic genotype overall and was significantly more prevalent among participants living with HIV than among HIV-negative individuals (44.7% vs. 21.2%, p=0.001).

### HPV Type Distribution by HIV Status

Distribution of HPV genotypes varied by HIV status (Figure 2; Supplementary Table 1). HPV33 was detected exclusively among HIV-positive participants, while HPV58 showed higher prevalence in this group, although the difference did not reach statistical significance. In contrast, HPV52 was significantly more common among HIV-negative individuals (11.3% vs. 3.5%, p=0.043). Among low-risk types, HPV 6/11 was significantly more prevalent among HIV-negative participants compared to those living with HIV (61.3% vs. 36.8%, p=0.001).

**Figure 1:**
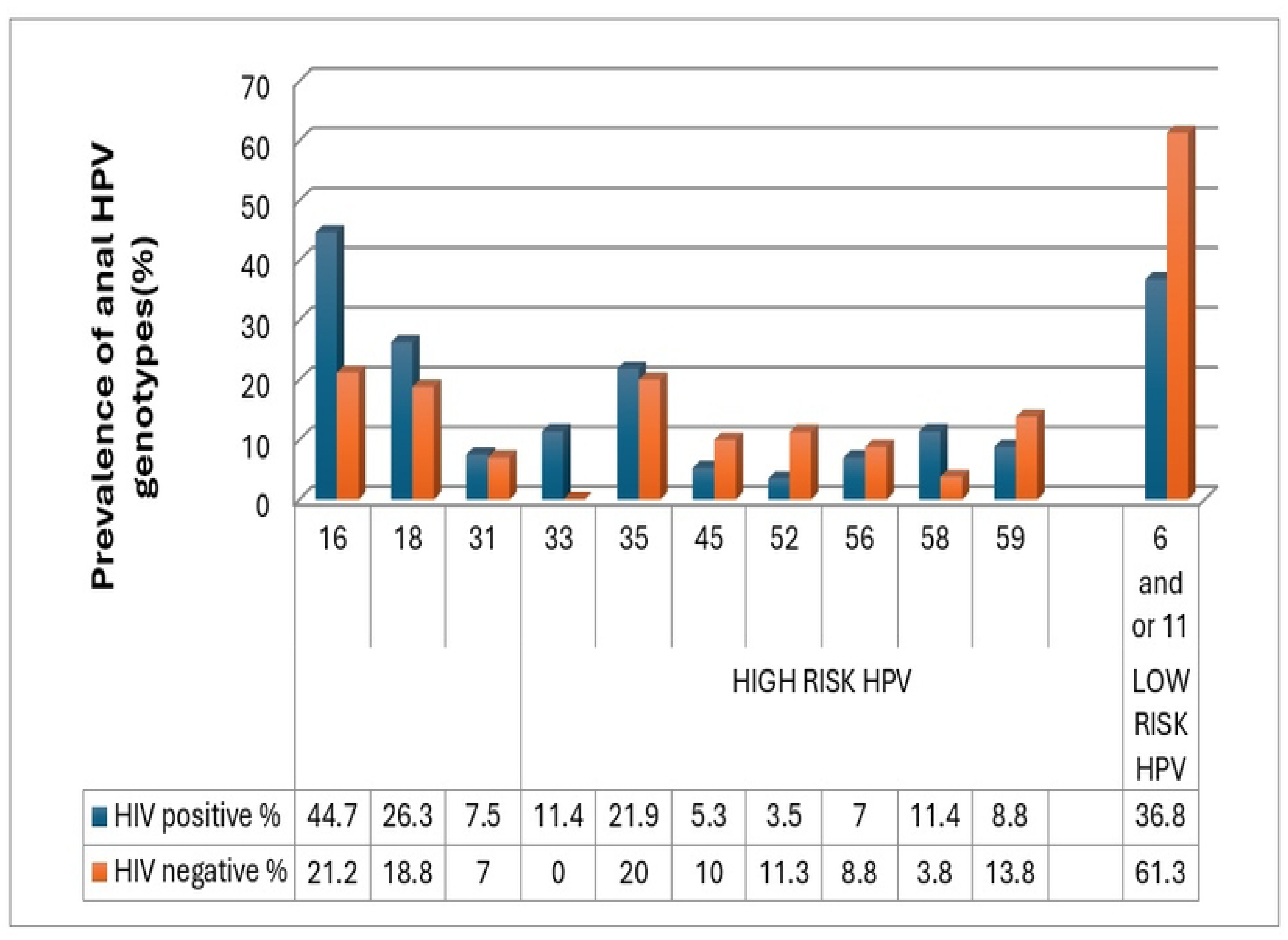
Prevalence of anal HPV genotypes by HIV status among MSM and transgender women

**Figure 2:**
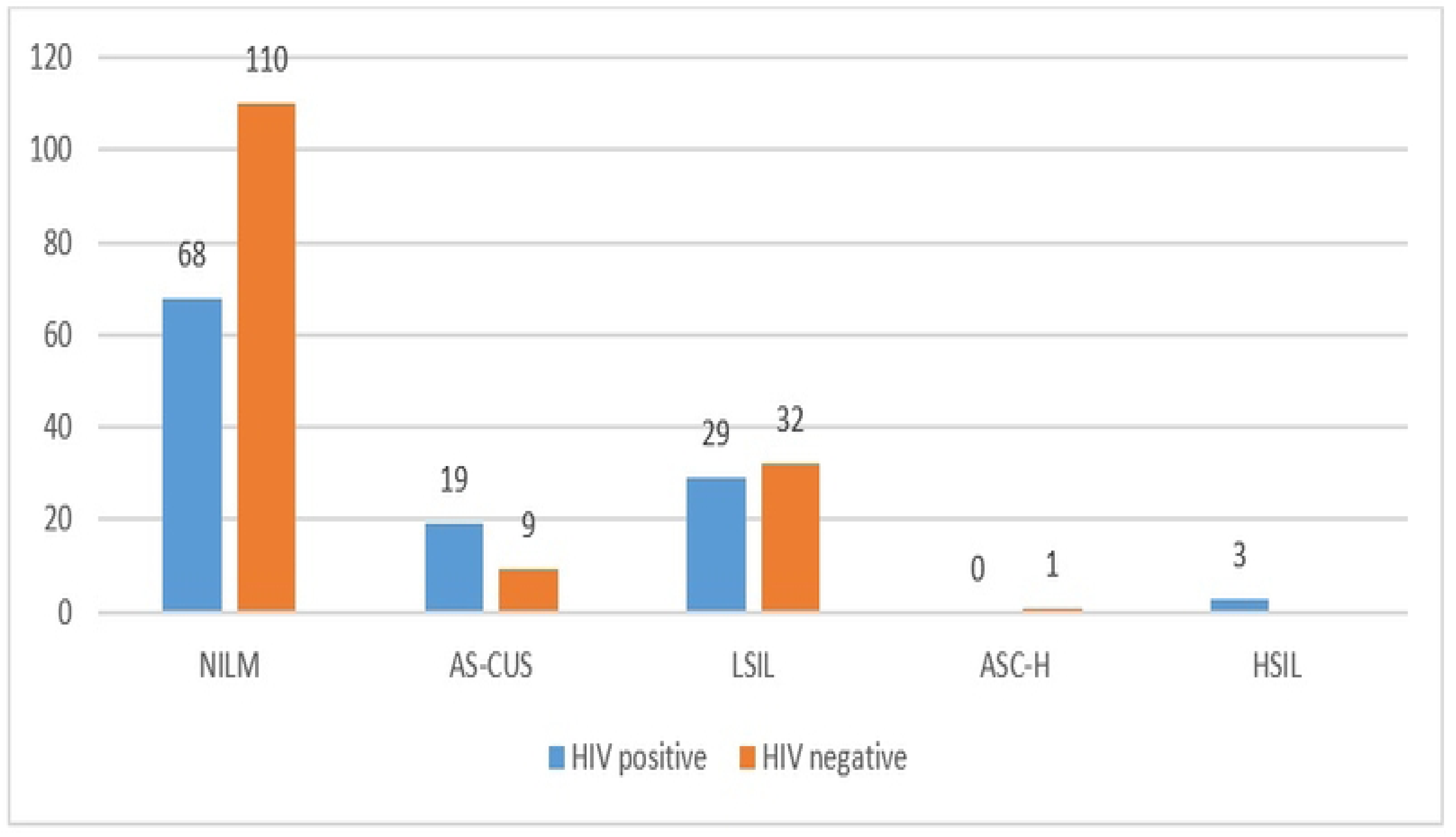
Distribution of Anal Squamous Intraepithelial Lesions (ASIL) stratified on HIV status among MSM and transgender women **NILM:** Negative for Intraepithelial Lesions or Malignancy **ASCUS:** Atypical Squamous Cells of Undetermined Significance **LSIL:** Low grade Squamous Intraepithelial Lesions **ASC-H:** Atypical Squamous Cells do not exclude high-grade lesion **HSILs:** High grade Squamous Intraepithelial Lesions

Bivalent vaccine-targeted HPV types (HPV16/18) were significantly more common among HIV-positive participants (65.8% vs. 40.0%, p<0.001), while no significant differences were observed for quadrivalent and nonavalent vaccine-targeted types.

### Characteristics of HIV–HPV Co-infected versus HPV-only Participants

Participants co-infected with HIV and HPV differed significantly from those with HPV infection alone across multiple demographic and behavioral characteristics (Table 2). Individuals in the co-infected group were older on average compared to HPV-only participants (mean age 29.9 vs. 25.4 years, p<0.001) and were more likely to be married (21.9% vs. 8.8%, p=0.034).

**Table 2:**
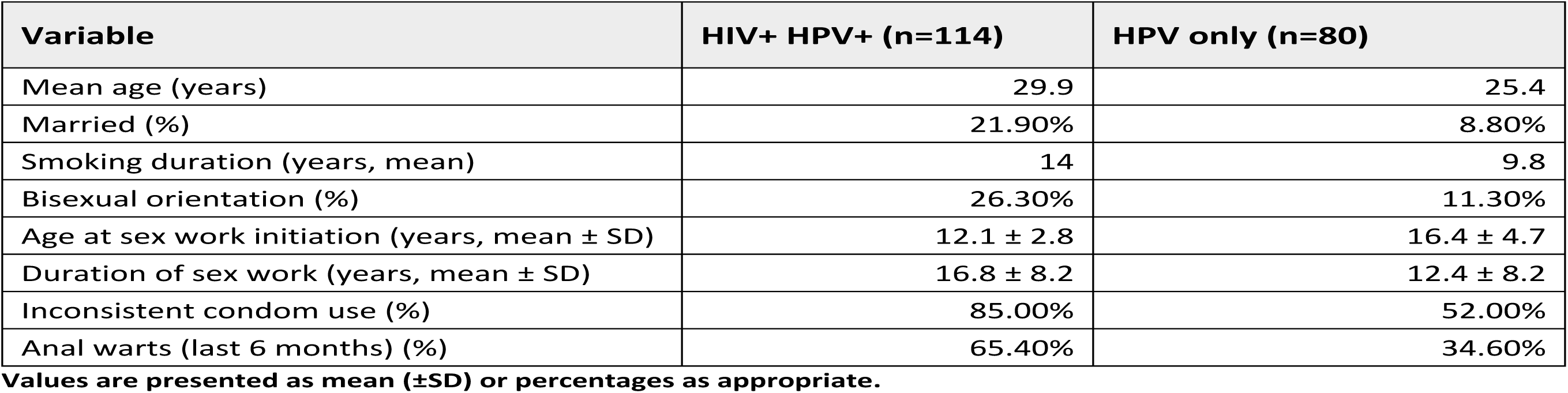
Characteristics of HIV–HPV Co-infected vs HPV-only Participants (n = 194)

| Variable | HIV+ HPV+ (n=114) | HPV only (n=80) |
| --- | --- | --- |
| Mean age (years) | 29.9 | 25.4 |
| Married (%) | 21.90% | 8.80% |
| Smoking duration (years, mean) | 14 | 9.8 |
| Bisexual orientation (%) | 26.30% | 11.30% |
| Age at sex work initiation (years, mean ± SD) | 12.1 ± 2.8 | 16.4 ± 4.7 |
| Duration of sex work (years, mean ± SD) | 16.8 ± 8.2 | 12.4 ± 8.2 |
| Inconsistent condom use (%) | 85.00% | 52.00% |
| Anal warts (last 6 months) (%) | 65.40% | 34.60% |

Values are presented as mean ( ± SD) or percentages as appropriate.

Behavioral risk profiles also differed markedly between the two groups. Co-infected participants reported a longer duration of smoking (mean 14.0 vs. 9.8 years, p=0.002) and were more likely to identify as bisexual (26.3% vs. 11.3%, p=0.010). Early initiation into sex work was more common among co-infected individuals, who reported a significantly lower age at entry (12.1 vs. 16.4 years, p=0.007) and a longer duration of engagement in sex work (16.8 vs. 12.4 years, p<0.001).

Sexual risk behaviors were also more pronounced among co-infected participants, with a substantially higher proportion reporting inconsistent condom use (85.0% vs. 52.0%, p<0.001).

In addition, recent anal warts were significantly more common in the co-infected group (65.4% vs. 34.6%, p=0.004), indicating a higher burden of concurrent sexually transmitted infections. (Table 2)

Overall, these findings suggest that HIV–HPV co-infection is concentrated among individuals with prolonged exposure to high-risk sexual networks, earlier entry into risk environments, and cumulative behavioral vulnerabilities. This clustering of demographic, behavioral, and clinical factors reflects the intersecting pathways through which co-infection may be sustained and amplified within this population.

### Prevalence of HPV by Sexual Orientation and HIV Status

HPV prevalence varied by sexual orientation within HIV strata. Among participants living with HIV, transgender women demonstrated the highest prevalence of both any HPV infection (96.9%) and high-risk HPV (81.8%), followed by homosexual men (88.1% and 74.5%, respectively) and bisexual men (76.9% and 51.3%) (p<0.001). (Table 3)

**Table 3:** Prevalence of any HPV and any high-risk HPV by HIV status and sexual orientation.

|  | HIV POSITIVE |  |  |  |  |  | HIV NEGATIVE |  |  |  |  |  |
| --- | --- | --- | --- | --- | --- | --- | --- | --- | --- | --- | --- | --- |
|  | OVERALL |  | Any HPV |  | Any High-Risk HPV |  | OVERALL |  | ANY HPV |  | Any High-Risk HPV |  |
|  | n | % | n | % | n | % | n | % | n | % | n | % |
| **HOMOSEXUALS** | 59 | 45.03 | 52 | 88.13 | 44 | 74.57 | 108 | 64.67 | 55 | 50.92 | 40 | 37.03 |
| **BISEXUALS** | 39 | 29.77 | 30 | 76.92 | 20 | 51.28 | 29 | 17.36 | 9 | 31.03 | 6 | 20.68 |
| **TRANSGENDER** | 33 | 25.20 | 32 | 96.96 | 27 | 81.81 | 30 | 17.96 | 16 | 53.33 | 13 | 43.33 |

A similar pattern was observed among HIV-negative participants, with transgender women again exhibiting higher prevalence of both any HPV infection (53.4%) and high-risk HPV (43.3%) compared to homosexual and bisexual men (p=0.001). (Table 3)

### Distribution of Anal Cytological Abnormalities by HIV Status

Among the 271 participants included in cytological analysis, 35.0% (n=93) had anal squamous intraepithelial lesions (ASIL). The prevalence of ASIL was significantly higher among individuals living with HIV compared to HIV-negative participants (42.4% vs. 28.1%, p≤0.001).

As shown in Figure 2, the distribution of cytological abnormalities varied by HIV status. Low-grade squamous intraepithelial lesions (LSIL) constituted the majority of abnormalities across both groups. However, higher-grade abnormalities were predominantly observed among individuals living with HIV. Notably, high-grade squamous intraepithelial lesions (HSIL) were detected only among HIV-positive participants, and all such cases were associated with HPV16 infection. In contrast, only a single HIV-negative participant presented with ASC-H, and no HSIL cases were observed in this group.

Participants without HIV were more likely to have normal cytology (NILM) or lower-grade abnormalities, indicating a comparatively lower burden of clinically significant lesions. Overall, these findings demonstrate a shift toward greater cytological severity among individuals living with HIV.

### Multiplicity of HPV Infection and Its Distribution by HIV Status and ASIL

Multiple HPV infections were significantly more common among individuals living with HIV compared to HIV-negative participants (p<0.001). Co-occurrence of multiple HPV genotypes was frequently observed among HIV-positive individuals. In addition, HIV seropositivity was 3.43 times more prevalent (Supplementary Table 2) among participants with high-risk HPV infection.

Multiplicity of HPV infection also varied by cytological status. Participants with anal squamous intraepithelial lesions (ASIL) were more likely to have multiple HPV types compared to those without ASIL (p<0.001). Among ASIL-positive individuals, a higher proportion had two or more HPV types, whereas participants without ASIL were more likely to have no detectable HPV or a single HPV type. (Table 4)

**Table 4.** Multiplicity of HPV types among any anal squamous intraepithelial lesions (ASIL) and HIV status among MSM and transgender women in Karachi Pakistan.

| Number of Types | All N=271<br>n (%) | ASIL Positive<br>n= 93<br>n (%) | ASIL Negative<br>n=178<br>n (%) | p-value | HIV Positive<br>n= 118<br>n (%) | HIV Negative<br>n= 153<br>n (%) | p-value |
| --- | --- | --- | --- | --- | --- | --- | --- |
| None | 92 (33.9) | 0 (13.0) | 92 (52.3) | <0.001* | 15 (12.7) | 77 (50.3) | <0.001 |
| One Type | 64 (25.1) | 32 (40.0) | 32 (17.0) |  | 27 (22.9) | 23 (15.1) |  |
| Two types | 84 (29.5) | 40 (37.9) | 44 (25.0) |  | 47 (39.8) | 35 (22.9) |  |
| Three types | 27 (10.0) | 20(21.1) | 07 (4.0) |  | 24 (20.3) | 12 (7.8) |  |
| Four types | 4 (1.5) | 1 (1.1) | 3 (1.7) |  | 5 (4.3) | 6 (3.9) |  |

P- value < 0.001 \*Fisher exact test p value

Stratified by HIV status, individuals living with HIV demonstrated a higher burden of multiple HPV infections across both ASIL-positive and ASIL-negative groups (p<0.001). Among participants with ASIL, higher numbers of HPV types were more frequently observed in HIV-positive individuals compared to HIV-negative individuals. (Table 4)

High-grade squamous intraepithelial lesions (HSIL) were observed only among individuals living with HIV, and all HSIL cases were positive for HPV16. In contrast, only one HIV-negative participant presented with ASC-H.

### Risk Factors Associated with HPV Infection and Anal Cytological Abnormalities

Multivariable analysis demonstrated that HIV infection was significantly associated with HPV infection, including HPV16. Participants living with HIV had a higher prevalence of any HPV infection (adjusted PR 1.44, 95% CI 1.05–1.97) and HPV16 infection (adjusted PR 2.81, 95% CI 2.16–3.82).

The presence of other sexually transmitted infections (STIs) was also associated with increased prevalence of HPV infection (adjusted PR 1.29, 95% CI 1.02–1.91) and showed a positive association with HPV16 infection, although this did not reach statistical significance.

Behavioral factors were associated with HPV infection. Inconsistent condom use (adjusted PR 2.01, 95% CI 1.07–5.20) and never using condoms (adjusted PR 1.58, 95% CI 1.03–4.44) were both associated with higher prevalence of HPV infection. Similar patterns were observed for HPV16 infection.

For anal cytological abnormalities, the presence of any high-risk HPV type was strongly associated with ASIL (adjusted PR 3.04, 95% CI 1.75–5.26). Co-infection with HIV and HPV was also associated with ASIL (adjusted PR 1.75, 95% CI 1.07–2.88), as was the presence of other STIs (adjusted PR 2.13, 95% CI 1.28–3.55). (Table 5) (Supplementary Table 3)

**Table 5:** Factors Associated with HPV Infection and Anal Cytological Abnormalities (ASIL) among MSM and Transgender Women in Karachi, Pakistan.

| Variables | HPV Infection<br>(Adjusted PR,<br>95% CI) | HPV16<br>Infection<br>(Adjusted PR,<br>95% CI) | ASIL (Adjusted PR,<br>95% CI) |
| --- | --- | --- | --- |
| HIV status (positive vs negative) | 1.44 (1.05–1.97) | 2.81 (2.16–3.82) | — |
| Any high-risk HPV | — | — | 3.04 (1.75–5.26) |
| Other STI (yes vs no) | 1.29 (1.02–1.91) | 1.52 (0.99–2.37) | 2.13 (1.28–3.55) |
| HIV–HPV co-infection | — | — | 1.75 (1.07–2.88) |
| Condom use<br>Inconsistent vs consistent<br>Never vs consistent | 2.01 (1.07–5.20)<br>1.58 (1.03–4.44) | 2.61 (1.50–4.50)<br>3.08 (1.69–5.60) | — |
| Preferred anal sex role (receptive<br>vs insertive) | 1.56 (0.87–2.83) | — | — |
| Smoking (yes vs no) | 1.17 (0.71–1.90) | — | — |

## DISCUSSION

This study provides robust evidence of an interrelated disease burden linking HIV infection, HPV acquisition, and anal squamous intraepithelial lesions (ASIL) among MSM and transgender women in Pakistan, highlighting overlapping biological and epidemiological vulnerabilities. The markedly elevated prevalence of HPV among individuals living with HIV, the concentration of high-risk HPV genotypes within this group, and the independent association between HIV–HPV co-infection and ASIL collectively indicate a strong relationship between these conditions and precancerous anal disease, consistent with findings from other MSM populations globally(3, 32). These findings extend existing evidence by situating these associations within a low-resource, high-stigma context where data have largely been absent (32).

The higher burden of HPV infection observed among individuals living with HIV aligns with well-established immunological mechanisms. HIV-induced immunosuppression compromises cell-mediated immunity, which is critical for HPV clearance, thereby increasing susceptibility to acquisition and persistence of infection(33–36). Persistent infection with oncogenic HPV types, particularly HPV16, has been identified as the central driver of anal carcinogenesis(37–39). In our study, the disproportionate clustering of high-risk HPV types and multiple HPV infections among HIV-positive participants reinforces this mechanism. The presence of multiple genotypes, which has been associated with viral persistence and synergistic oncogenic effects, further supports the hypothesis that HIV infection creates a biological environment conducive to HPV progression rather than mere acquisition(39–42).

Importantly, the finding that all participants with ASIL were HPV-positive provides compelling empirical support for a stepwise biological progression from infection to dysplasia. This observation is consistent with prior studies demonstrating that HPV infection is a necessary precursor for anal intraepithelial neoplasia and subsequent malignancy(43, 44). Moreover, the exclusive occurrence of high-grade lesions among individuals living with HIV in our study underscores the role of immune status in modulating disease severity. Although antiretroviral therapy (ART) improves immune function and survival, evidence suggests that it does not fully mitigate the risk of HPV persistence or progression to anal dysplasia, likely due to incomplete immune reconstitution at mucosal sites(45, 46). Our findings are consistent with this body of work and highlight the need for targeted HPV-related interventions even among individuals receiving ART.

A key contribution of this study lies in disentangling the roles of biological and behavioral determinants. While behavioral factors such as number of sexual partners, condom use, and duration of sex work are well-recognized drivers of exposure, they did not retain statistical significance in adjusted models once biological variables were included. This does not diminish their importance; rather, it suggests that their effects are mediated through biological pathways, principally HPV acquisition and co-infection. For instance, condom use, although protective against many sexually transmitted infections, provides limited protection against HPV due to its transmission through skin-to-skin contact(47). Similarly, high partner turnover increases exposure probability but does not independently predict cytological abnormalities when HPV infection is already established. This distinction between upstream exposure factors and proximal biological drivers is critical for both interpretation and intervention design.

These findings are further supported by multivariable analysis, which demonstrated that biological factors, rather than behavioral exposures alone, were the primary determinants of both HPV infection and progression to anal dysplasia. HIV infection was independently associated with a higher prevalence of HPV infection, particularly HPV16, reinforcing its role as a key upstream driver of oncogenic HPV acquisition. In addition, the presence of high-risk HPV types was the strongest predictor of ASIL, while co-infection with HIV and HPV and the presence of other sexually transmitted infections (STIs) remained independently associated with cytological abnormalities. Although behavioral factors such as inconsistent condom use showed associations with HPV infection, they did not retain significance in adjusted models for ASIL, suggesting that their effects are mediated through biological pathways rather than acting as direct determinants of disease progression. Together, these findings support a hierarchical model in which behavioral exposures increase the likelihood of HPV acquisition, while biological factors, particularly HIV-associated immune dysfunction and co-infection with oncogenic HPV types, drive persistence and progression to anal dysplasia.

Moreover, the strong association between concurrent sexually transmitted infections (STIs) and ASIL observed in this study further supports a syndemic framework. Co-occurring infections may act synergistically by inducing local inflammation, disrupting epithelial integrity, and facilitating viral persistence and integration(48, 49). In this context, HIV, HPV, and other STIs do not operate in isolation but interact within a network of biological and social vulnerabilities, amplifying disease burden.

These biological interactions must also be interpreted in light of the rapidly evolving HIV epidemic in Pakistan. Recent evidence indicates a substantial increase in HIV infections over the past few years, with a large proportion of individuals remaining undiagnosed and ongoing transmission concentrated within key populations, including MSM and transgender women. This expanding epidemic creates conditions for sustained exposure to HPV and other sexually transmitted infections, thereby amplifying the risk of co-infection and progression to anal dysplasia. In this context, the observed clustering of high-risk HPV and HIV in our study reflects a broader epidemiological shift, in which an increasing HIV burden may act as a catalyst for HPV-related disease progression. These biological interactions must be further interpreted within the broader structural context of Pakistan, where MSM and transgender women experience significant stigma, marginalization, and barriers to healthcare access. Limited availability of targeted sexual health services, absence of routine HPV screening, and exclusion of HPV vaccination from the national immunization program contribute to delayed diagnosis and untreated infections(50). The high prevalence of sex work and dense sexual networks observed in this study are not merely individual risk behaviors but reflect constrained socioeconomic conditions and limited livelihood options. Such structural vulnerabilities create conditions in which both HIV and HPV transmission are sustained and amplified, consistent with syndemic theory (51, 52).

From a public health perspective, these findings have important implications. The demonstrated biological continuum from HIV infection to HPV-related dysplasia highlights the need for integrated prevention strategies. Incorporating HPV screening into existing HIV care programs could facilitate early detection of precancerous lesions, while targeted HPV vaccination for high-risk populations may reduce future disease burden(28). Evidence from other settings suggests that anal cytology screening, although not universally implemented, can be an effective and cost- efficient strategy for high-risk groups(53, 54). In Pakistan, where both HPV vaccination and anal cancer screening are currently limited, such integrated approaches are particularly urgent.

This study has several strengths, including the use of both molecular and cytological methods, the inclusion of HIV-positive and HIV-negative participants, and the generation of context-specific evidence from a previously understudied population. However, the cross-sectional design limits causal inference, and the absence of high-resolution anoscopy precludes histological confirmation of lesions. Despite these limitations, the consistency of findings with established biological mechanisms and global literature supports the validity of the observed associations.

In conclusion, this study demonstrates a clear syndemic interaction between HIV and HPV among MSM and transgender women in Pakistan, characterized by increased HPV prevalence, multiplicity of infection, and progression to anal dysplasia. The findings support a biological continuum model in which HIV infection facilitates HPV persistence and accelerates disease progression. In the context of a rapidly evolving HIV epidemic in Pakistan, with ongoing transmission concentrated within key populations and substantial gaps in diagnosis and treatment, this co-infection dynamic represents an emerging and under-recognized pathway to HPV-related disease burden. Addressing this dual burden will require integrated interventions that move beyond single-disease approaches to simultaneously target biological co-infection, behavioral exposures, and structural barriers to care, including the incorporation of HPV prevention and screening within existing HIV programs.

## IMPLICATIONS AND RECOMMENDATIONS

In the context of a rapidly evolving HIV epidemic in Pakistan, the intersection of HIV and HPV represents an emerging pathway to increased HPV-related disease burden, particularly among key populations such as MSM and transgender women. The findings of this study therefore carry important implications for public health policy, clinical practice, and future research in Pakistan and similar low- and middle-income settings. The demonstrated biological continuum linking HIV infection, HPV co-infection, and progression to anal dysplasia underscores the need to move beyond disease-specific approaches toward coordinated models of prevention and care.

At the policy level, the high burden of HPV infection and its concentration among individuals living with HIV highlight a critical gap in the current national response. Despite growing global recognition of HPV as a major contributor to cancer burden, HPV vaccination is not yet incorporated into Pakistan’s national immunization program. These findings support prioritizing HPV vaccination not only for adolescent girls as part of cervical cancer prevention, but also for high-risk populations such as MSM and transgender women. Targeted vaccination strategies, particularly those integrated within existing HIV programs, may offer a feasible and cost-effective approach in resource-constrained settings.

From a health systems perspective, the strong association between HIV-HPV co-infection and anal dysplasia calls for incorporation of HPV-related services within HIV care platforms. Routine HIV programs in Pakistan provide an existing infrastructure that can be leveraged to incorporate HPV screening, early detection of anal lesions, and referral pathways for further management. Given the high prevalence of anal squamous intraepithelial lesions observed in this study, introducing anal cytology screening for high-risk groups may enable earlier identification of precancerous conditions and reduce progression to malignancy. While high-resolution anoscopy may not be widely available, phased implementation of simpler screening approaches could serve as an important entry point.

The observed association between concurrent STIs and anal dysplasia further underscores the need to strengthen sexually transmitted infection services. Integrated STI diagnosis and treatment may help interrupt the biological processes that facilitate HPV persistence and progression. Expanding access to comprehensive sexual health services, including syndromic management and laboratory-based diagnostics, will be essential in addressing the broader syndemic of infections affecting these populations.

At the community level, interventions must address the structural and social barriers that shape vulnerability to both HIV and HPV. Stigma, discrimination, and limited access to healthcare services remain significant obstacles for MSM and transgender women in Pakistan. Community-based approaches that engage peer networks, improve health literacy, and build trust in healthcare systems are critical for increasing uptake of prevention and screening services. Without addressing these underlying social determinants, biomedical interventions alone are unlikely to achieve sustained impact.

The findings also carry important implications for research. Longitudinal studies are needed to further elucidate the temporal progression from HPV infection to anal dysplasia and cancer in this population. Additionally, implementation research is required to evaluate the feasibility, acceptability, and cost-effectiveness of integrating HPV vaccination and screening within existing HIV programs. Generating local evidence on these interventions will be essential for informing policy decisions and scaling up effective strategies.

In conclusion, addressing the dual burden of HIV and HPV in Pakistan requires a coordinated, multi-level response integrating vaccination, screening, clinical care, and community engagement. In the context of an expanding HIV epidemic, such approaches will be critical for reducing HPV-related disease burden and improving long-term health outcomes among marginalized populations.

## Authors contribution

Alyan Ahmed conceptualized the manuscript, initiated the analysis, conducted the descriptive analyses, and led the initial manuscript drafting. Shaheer Hasan Rizvi, Ali Ahmed Rizvi, and Fabeeha Ali contributed to data analysis, interpretation, and manuscript writing. Aymen Haroon reviewed multiple manuscript versions and contributed to manuscript refinement. Muslima Ejaz along with Alyan Ahmed conceptualizes the manuscript, provided overall academic supervision, critical review and interpretation of the analyses, continuous mentorship throughout the analytical and writing process, and substantial intellectual input to manuscript development. All authors reviewed and approved the final manuscript.

## Data availability

The data supporting the findings of the original research project are available from the Aga Khan University Hospital Karachi Pakistan, Department of Community Health Sciences, and may be accessed with permission from corresponding author.

## Patient and public Involvement

Patients and/or public were not involved in the design, or conduct, or reporting, or dissemination plans of this research

## Patient consent for publication

Not applicable

## Ethical approval

The Ethics committee on Human Research of Aga Khan (3612-CHS-ERC-15) and Dow (IRB-557/DUHS/APPROVAL/2015/84) Universities of Health sciences approved the study

